# SARS2 simplified scores to estimate risk of hospitalization and death among patients with COVID-19

**DOI:** 10.1101/2020.09.11.20190520

**Authors:** Hesam Dashti, Elise C. Roche, David William Bates, Samia Mora, Olga Demler

## Abstract

Although models have been developed for predicting severity of COVID-19 based on the medical history of patients, simplified risk prediction models with good accuracy could be more practical. In this study, we examined utility of simpler models for estimating risk of hospitalization of patients with COVID-19 and mortality of these patients based on demographic characteristics (sex, age, race, median household income based on zip code) and smoking status of 12,347 patients who tested positive at Mass General Brigham centers. The corresponding electronic health records were queried from 02/26/2020 to 07/14/2020 to construct derivation and validation cohorts. The derivation cohort was used to fit a generalized linear model for estimating risk of hospitalization within 30 days of COVID-19 diagnosis and mortality within approximately 3 months for the hospitalized patients. On the validation cohort, the model resulted in c-statistics of 0.77 [95% CI: 0.73-0.80] for hospitalization outcome, and 0.72 [95% CI: 0.69-0.74] for mortality among hospitalized patients. Higher risk was associated with older age, male sex, black ethnicity, lower socioeconomic status, and current/past smoking status. The model can be applied to predict risk of hospitalization and mortality, and could aid decision making when detailed medical history of patients is not easily available.

## Introduction

On 29 August 2020, the Centers for Disease Control and Prevention (CDC) reported 291,985 new COVID-19 weekly cases in the U.S. that increased the total number of cases in the U.S. to 5,890,532 patients^1^. At the rise of the new surge in cases, designing models for predicting severity of COVID-19 illness is essential for public health strategies, as risk scores could enable allocations of limited medical resources and preparedness of healthcare facilities. The CDC reports age and medical comorbidities (e.g. chronic kidney disease, heart conditions, immunocompromised conditions, obesity, etc.) as leading risk factors of severe illness in patients with COVID-19^2^. The importance of these risk markers has been studied^3–16^, and significance of associations between severity of illness and different patient characteristics have been demonstrated. These studies reported association between higher age and severe illness, prepandemic health disparities and higher risk of severe COVID-19 outcomes in blacks and racial minorities^9,10,17^, importance of obesity^18^ and its impacts on infected children and adults^8,19,20^, increased severity of COVID-19 illness in immunodeficient patients^4,11^, the role of preexisting cardiovascular disease (CVD) and the use of cardiovascular medications^21–24^ on severity of outcomes, and effects of kidney and pulmonary diseases^3^. Smoking has also been associated with COVID-19 outcomes^25–28^. The largest COVID-19 cohort study on more than 10,000 COVID-19 related deaths in the U.K.^12^ indicated a few preexisting medical conditions were significantly associated with severity in non-white and low socioeconomical regions. In another study on mortality of patients with COVID-19 in intensive care units (ICU) in the Lombardy region of Italy, older age, male sex, and measured arterial oxygenation parameters on admission to ICU were independently associated with mortality, while they also identified risk factors from patients’ medical history (chronic obstructive pulmonary disease, hypercholesterolemia, and type 2 diabetes)^5^. In a similar study in the U.S., mortality rate of ICU patients was associated to older age, male sex, high body mass index, arterial oxygenation, liver and kidney disfunction on admission, and medical history of coronary artery disease and active cancer were independently associated with mortality^7^.

In these studies, the list of investigated and recorded risk markers from medical history of patients varied, which could be due to the complexity and challenges associated with extracting phenotypes from electronic health records (EHR) data^29–32^. Hence, a simplified model that can accurately predict severity of the illness without the need of detailed examination of medical history could be more practical. In addition, patient characteristics on admission have been demonstrated to be strongly associated with the severity of illness, and the most common risk markers have been demographic variables. Therefore, we hypothesized that a simplified risk score may provide a fast and reliable prediction of hospitalization of patients with COVID-19 and mortality among these patients. We examined this hypothesis using demographic variables and smoking status of patients tested positive for COVID-19 at Mass General Brigham (MGB) medical centers, Massachusetts, U.S.A.

## Results

The examined population contained N = 12,347 patients tested positive for COVID-19 at MGB facilities. This population consists of 42.77% white, 15.91% black, 9.05% Hispanic, and 32.28% other/unknown races. Cumulative endpoints were 3,401 hospitalized patients, from which 509 were deceased. Characteristics of these patients are shown in Table 1.

**Table 1.** Characteristics of N=12347 patients with COVID-19 from the Mass General Brigham electronic health records.

|  |  |  |  |  |
| --- | --- | --- | --- | --- |
| **Characteristics** | **Outpatients**<br>**N=8946** | **Hospitalized**<br>**N=3401** | **Deceased**<br>**inpatients**<br>**N=509** | **Total**<br>**N=12347** |
| Age (year), Median (IQR) | 42.0 (29.0-56.0) | 62.0 (48.0-77.0) | 78.0 (69.0-87.0) | 47.0 (32.0-62.0) |
| Women, N (%) | 4969 (55.5) | 1616 (47.5) | 215 (42.2) | 6585 (53.3) |
| Race, N (%) |  |  |  |  |
| White | 3564 (39.8) | 1717 (50.5) | 322 (63.3) | 5281 (42.8) |
| Black | 1371 (15.3) | 593 (17.4) | 79 (15.5) | 1964 (15.9) |
| Hispanic | 920 (10.3) | 197 (5.8) | 27 (5.3) | 1117 (9.0) |
| Other/Not recorded | 3091 (34.6) | 894 (26.3) | 81 (15.9) | 3985 (32.3) |
| Median household income (\$1000), Median (IQR) | 60.4 (53.3-86.2) | 65.5 (53.3-94.8) | 68.3 (55.0-98.6) | 64.2 (53.3-90.5) |
| Smoking, N (%) |  |  |  |  |
| Current | 353 (3.9) | 210 (6.2) | 20 (3.9) | 563 (4.6) |
| Former | 1099 (12.3) | 860 (25.3) | 171 (33.6) | 1959 (15.9) |
| Never | 5133 (57.4) | 1920 (56.5) | 164 (32.2) | 7053 (57.1) |
| Unknown | 2361 (26.4) | 411 (12.1) | 154 (30.3) | 2772 (22.5) |

IQR: interquartile range

### Predicting risk of hospitalization

The fitted generalized linear model (GLM) in the derivation cohort of MGB’s non-employees (N = 10,496, 30.46% hospitalized) indicated significant associations between the examined variables and hospitalization (Table 2). The odds ratios (OR) indicated higher risks of hospitalization for older and male patients. Compared with white patients, Hispanic patients had lower risk of hospitalization while black patients were at the highest risk (test of trend p-value < 0.001). Although the OR of median household income was close to 1, higher income was associated with lower risk of hospitalization. Test for trend in smoking status was significant (p-value < 0.001) with current smokers at the highest risk, followed by former smokers, and finally non-smokers at a lower risk of hospitalization.

**Table 2.** Adjusted odds ratios of the examined variables for predicting risk of hospitalization among patients with COVID-19 (N=10,496).

| Variables | Median (IQR), N (%) | OR (95% CI) | P |
| --- | --- | --- | --- |
| Age (years) | 48.0 (32.0-64.0) | 1.53 (1.49-1.57) | <.001 |
| Sex |  |  |  |
| Female | 5251 (50.03) | ref | ref |
| Male | 5245 (49.97) | 1.40 (1.28-1.54) | <.001 |
| Race |  |  | <.001* |
| White | 4444 (42.34) | ref | ref |
| Black | 1472 (14.02) | 1.30 (1.13-1.49) | <.001 |
| Hispanic | 974 (9.28) | 0.58 (0.48-0.70) | <.001 |
| Other/Not recorded | 3606 (34.36) | 1.02 (0.91-1.15) | 0.74 |
| Median household income (\$1000) | 60.4 (53.3-86.2) | 0.98 (0.96-0.99) | 0.007 |
| Smoking |  |  | <.001* |
| Current | 489 (4.66) | 1.44 (1.17-1.76) | <.001 |
| Former | 1772 (16.88) | 1.22 (1.08-1.38) | 0.002 |
| Never | 5715 (54.45) | ref | ref |
| Unknown | 2520 (24.01) | 0.53 (0.46-0.60) | <.001 |

Each variable shown was mutually adjusted for the other variables in the table. IQR: interquartile range. Medians, interquartile ratios, and percentages are reported on the derivation population. Odd ratios (OR) and the corresponding 95% confidence interval (CI) for age is reported per 10 years increment, and these values for median household income are shown per 10,000\$.
\* Test of trend p-value.

Examining this model in the validation cohort of MGB employees (N=1,851, 11.02% hospitalized) showed an area under the curve (AUC) of 0.77 [95% CI: 0.73-0.80] (Supplementary Figure 2a). The optimal predicted probability cutoff for discriminating between the two groups was 0.29, and the second optimal cutoff for identifying an intermediate risk group was 0.16. After applying these cutoffs on the MGB employees, the resulting receiver operating characteristic curve had an AUC of 0.73 [95% CI: 0.70-0.76]. The model is well-calibrated in the validation cohort, based on the Hosmer-Lemeshow goodness of fit (GOF) test, p-value of 0.11. The GOF test was conducted after performing recalibration to adjust for different event rates in the derivation and validation cohorts. The corresponding calibration plot is shown in Supplementary Figure 2b. After categorizing age (0-29, 30-59, 60-79, >= 80; years) and median household income (<60, 60-80, >=80; $1000), a GLM was fit on the derivation cohort and the model performed consistently with the main model (AUC in validation set: 0.75 [95% CI: 0.71-0.78]). The ORs of this model were consistent with the main model (Supplementary Table 2). Heatmap of risk scores according to this categorization of age and median income is presented in Supplementary Figure 1.

### Predicting mortality

The GLM model (Table 2) was then applied to predict death among hospitalized patients with COVID-19 (N=3,401, 14.97% deceased). The AUC was 0.72 [95% CI: 0.69-0.74] (Supplementary Figure 2c). The optimal predicted probability cutoff point for distinguishing deceased vs. alive hospitalized patients was 0.50, and the second cutoff was 0.28. Applying these cutoffs resulted in AUC of 0.70 [95% CI: 0.68-0.72]. Based on the Hosmer-Lemeshow GOF test, the model is well calibrated, p-value of 0.62 (Supplementary Figure 2d). An average AUC of 0.73 [95% CI: 0.72-0.73] was reached when the model was validated five times on 680 randomly selected hospitalized patients.

### Sensitivity analyses

Effects of MGB’s change of policies in COVID-19 testing criteria before and after April 29, 2020 were considered. Two GLM models were trained on MGB non-employees who were tested for COVID-19 before (N=6,624, 33.57% hospitalized) and after (N = 3,872, 25.13% hospitalized) April 29, 2020 that showed similar trends to the main model (Supplementary Table 3). Although the OR for median household incomes remained close to 1, the corresponding OR of the after April 29^th^ cohort showed a different direction (OR 1.04 [95% CI: 1.01-1.07], p-value 0.005) compared to the main model (OR 0.98 [95% CI: 0.96-0.99], p-value 0.007). The ORs of the other characteristics (age, sex, race, and smoking) from the main model were confirmed in both before and after cohorts. We examined performance of the main model for predicting mortality of hospitalized patients with reference COVID-19 date before(N = 2,379, 16.98% deceased) and after (N=1,022, 10.27% deceased) April 29^th^ (results not shown here). This analysis showed a good AUC of 0.73 [95% CI: 0.70-0.76] for the former group, and for the patients tested after April 29^th^, the model showed an AUC of 0.67 [95% CI:0.62-0.71]. In additional sensitivity analysis without excluding 24 patients who have been hospitalized after the 30 days interval, the model fit on MGB non-employees produced similar OR as the main model (results are not shown here). Validating this model on the MGB employee cohort produced similar AUC and confidence interval as the main model. An additional GLM was trained on MGB non-employees older than 60 years (N = 3,119, 55.59% hospitalized) to further investigate the lower rate of hospitalization among Hispanic patients compared to white and black patients. This population contained 1,878 white patients, 168 Hispanic, 459 black, and 614 other/unknown races, from which 57.51%, 41.07%, 60.35%, 50.16% were hospitalized, respectively. The model for this sensitivity analysis showed similar trends of odds ratios compared to the main model (not shown here), and resulted in an AUC of 0.69 [95% CI: 0.62-0.76]) on the corresponding validation cohort of MGB employees older than 60 years (N = 245, 33.47% hospitalized).

### Risk groups

The optimal predicted probability cutoffs for the model when used to estimate risk of hospitalization of patients with COVID-19 (0.29 and 0.16), and when the model was applied for predicting mortality among hospitalized patients (0.50 and 0.28) were used to define low, intermediate, and high-risk groups. The beta coefficients of the model were mapped according to 1 unit change that rescaled risk scores to 0-68 (Table 3). The rescaled cutoffs indicated high risk of hospitalization for patients with score ≥ 21, intermediate risk (9 ≤ score < 21), and low risk (score < 9). Similarly, high risk of mortality among hospitalized patients was assigned to scores ≥ 40, intermediate risk to 20 ≤ score < 40, and low risk patients have a score of less than 20.

**Table 3.** SARS2 risk scores.

| Characteristics |  |  | Score |
| --- | --- | --- | --- |
| Sex |  |  |  |
|  | Female |  | 0 |
|  | Male |  | +7 |
| Age, years |  |  |  |
|  | 0-29 |  | 0 |
|  | 30-59 |  | +1 |
|  | 60-79 |  | +24 |
| | $\geq 80$ | | +35 |
| Race |  |  |  |
|  | Hispanic |  | 0 |
|  | White/Other |  | +11 |
|  | Black |  | +16 |
| Socioeconomic status (median household income) |  |  |  |
| | $< \$60K$ | | +3 |
| | $\$60K-\$80K$ | | +1 |
| | $\geq \$80K$ | | 0 |
| Smoking status |  |  |  |
|  | Current |  | +7 |
|  | Ever |  | +4 |
|  | Never |  | 0 |

Hospitalization: Low risk: score $< 9$ , Intermediate risk: $9 \leq \text{score} < 21$ , High risk: score $\geq 21$ . Mortality among hospitalized patients: Low risk: score $< 20$ , Intermediate risk: $20 \leq \text{score} < 40$ , High risk: score $\geq 40$ .

## Discussion

Currently, the U.S. is one of the epicenters of the pandemic with an increasing number of COVID-19 cases and mortality. The capability of predicting severity of COVID-19 illness in a fast and efficient manner would help healthcare workers to distinguish high risk patients. We utilized MGB EHR data of patients with COVID-19 to design simplified models for predicting hospitalization risk and also risk of mortality among hospitalized patients, where the model requires only demographic variables (age, sex, race, median household income) and smoking status of the patients. Testing the models on the validation cohorts showed high AUC (0.77 and 0.72 for hospitalization and mortality), and applying discrimination cutoffs for distinguishing patients with severe illness resulted in good AUCs as well. The Hosmer-Lemeshow GOF test resulted in p-values > 0.05 indicating good calibration of the SARS2 model.

Model performance characteristics such as AUC and Hosmer-Lemeshow GOF test calculated in set-aside validation cohorts indicated that the model has good discrimination and calibration, and performed well in the population of MGB patients. The odds ratios reported for our model are consistent with the currently available knowledge about association of severity of COVID-19 with demographic characteristics. This model is named “SARS2”, for its input variables: Sex, Age, Race, Socioeconomics status, Smoking status. The proposed SARS2 model is provided as a web interface for seamless calculation of the risk scores and risk categories (https://dashti.bwh.harvard.edu/sars2/).

In the main and the sensitivity analyses, Hispanic patients had a lower risk compared to white and black patients. Although these results align with the lower rate of hospitalized Hispanic patients in the current CDC reports (Hispanic: 22.9%, white: 31.7%, and black: 32.9%)^33^, analysis on the MGB’s EHR records showed 84.33% of Hispanic patients with COVID-19 are younger than 60 years. The younger age could explain the lower rate of hospitalization, and further investigations on Hispanic patients are needed. The derivation and validation cohorts are from patients tested positive for COVID-19 at MGB medical centers, and further validation of the models on other cohorts is required to establish generalizability beyond our data. Because of the complexity of EHR data, admission diagnoses and causes of death were not considered in this study. Therefore, although non-COVID-19 related admission rates dropped during the pandemic, some of our hospitalization and mortality endpoints may not be due to COVID-19 illness.

The proposed SARS2 model for predicting hospitalization among COVID-19 patients, and mortality among hospitalized patients is designed based on easily accessible risk markers (age, sex, race, median household income, and smoking status). It is well known that extraction of a valid history of medication-use, and diagnoses and preconditions is not always feasible. Therefore, designing simplified models that can be used as prescreening at clinics increases the practicality and efficiency of these models in healthcare facilities. Although there is a limited number of risk scores available for predicting hospitalization or death among patients with COVID-19, the simple model presented here is on par with the c-statistics of more comprehensive models that for example predict mortality in the largest available COVID-19 cohort (average AUC of 0.77)^12^, or the survival model developed using cytokines, demographics and comorbidities on patients admitted to the Mount Sinai Health System in New York (AUC ranged from 0.65-0.76)^16^. The provided web interface for calculating risk scores enables easy assessment of risk of hospitalization and mortality proposed in this study.

## Methods

### Study Population

On 07/14/2020 a total of 12,460 individuals (outpatients and inpatients) have been diagnosed with COVID-19 at MGB medical centers. Demographic variables (age, sex, race, zip code), smoking status, hospital admission records, and COVID-19 lab results of these patients were queried from MGB’s EHR (Figure 1), and the institutional review board (IRB) approved this investigation of the EHR data. The COVID-19 lab results were dated within 03/04/2019-06/29/2020, and during this period, MGB employees working onsite underwent constant self-monitoring for symptoms and selective COVID-19 testing. The criteria for testing non-employees varied during the examined time interval; before April 29, 2020 symptomatic patients who were defined as high risk (e.g., age> =70, severe chronic lung disease, sever heart disease, on immunocompromising medications, reside in counties with high number of cases) or of specific categories (e.g., pregnant > = 36 weeks, patients being discharged) were tested. However, a more relaxed criteria were applied after April 29, 2020 such that testing was not dependent on older age or preexisting medical conditions, and instead the criteria were defined based on symptoms (e.g., documented fever, cough, anosmia).

**Figure 1.**
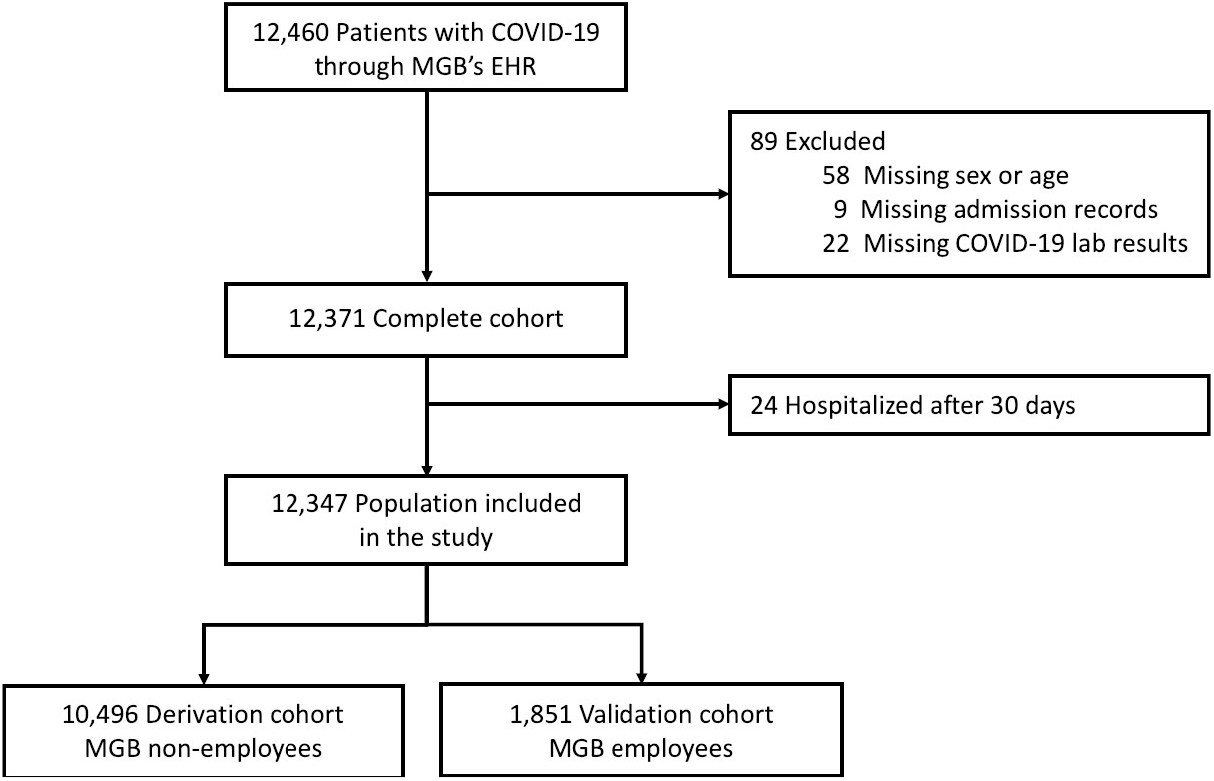
Study population diagram.

For every patient, the earliest positive (positive or presumptive positive) result of their COVID-19 tests was used as a reference date, and the time interval from these reference dates to the time of retrieving data for this study (07/14/2020) has a median follow-up of 84 days [95% IQR: 69-96 days]. The EHR contained patients labeled as COVID-19 positive when their lab test results were positive/presumptive positive or patients were diagnosed with COVID-19 infection by the medical staff at MGB centers (COVID-19 ICD codes were used). Those without available COVID-19 lab test results were excluded from this study. The deceased flag and its corresponding date were retrieved from the EHR that indicated date of death among hospitalized patients within 74 days from the date of COVID-19 diagnosis (median date of death: 9 days [95% IQR: 4-16 days]). Because of the waiting periods for receiving results of COVID-19 tests, hospital admission records dated between 7 days before until 30 days after patients reference date were queried from the EHR to identify hospitalized patients. Time to hospitalization ranged from −7 to 29 days with median of 0 days, that reflects a positive COVID-19 diagnosis was a requirement for hospitalization in most cases. We note that the examined patient characteristics (age, sex, race, zip code, smoking status) are independent from time of events (hospitalization or death), and an ideal testing condition, with immediate availability of results, will not change associations between the examined characteristics and the events. Therefore, the events are considered as cumulative endpoints for the examined follow-up duration. We verified that outpatients had no record of admissions (more than 2 days) to MGB medical facilities during the period of −7 to 30 days of follow-up.

In order to expand applications of the SARS2 model to more diverse regions in the U.S., we mapped patients’ primary zip codes to their median household incomes according to the U.S. Census 2018 data. These median household incomes were used as indicators of socioeconomic status of the patients. The EHR population contained 385 Asian, 18 Hawaiian, 30 American Indian, and 5 Dominicans that were considered as other races in the analysis.

MGB employees (validation cohort) and non-employees (derivation cohort) differed in their demographic characteristics (Supplementary Table 1) and also followed different COVID-19 testing criteria in the limited capacity setting. Presence of these differences between derivation and validation cohorts protects against over-optimism in estimating model performance characteristics and ensures robustness of the model. A logistic regression model (a generalized linear model with logit link (GLM)) was fit to predict hospitalization outcome. The same model was used for predicting mortality among the hospitalized patients.

To derive a model for predicting hospitalization of patients, we trained a GLM on demographic characteristics (sex, age, race, median household income), and smoking among non-employees (N = 10,496, 30.46% hospitalized) and validated the model on MGB employees (N= 1,851, 11.02% hospitalized). Because mortality was recorded for inpatients, we examined the model performance for estimating mortality of the hospitalized patients (N=3,401, 14.97% deceased). In addition, because of the relatively lower rates of mortality among MGB employees, an average c-statistics of 5 iterations of validating the prediction model on randomly selected 20% of the hospitalized patients was also reported.

### Statistical Methods

The EHR data were preprocessed using Python scripts. All variables (sex, age, race, median household income, and smoking status) were used in the R glm function to derive a multivariable model for predicting risk of hospitalization. In this model, linear associations with binomial distribution (logit link function) was used to distinguish between hospitalized vs. outpatient. The default glm convergence criteria on deviances was used to stop the iterations. The DeLong method was used to calculate confidence intervals for the c-statistics. The R coords function with Youden’s ‘best’ method was used to calculate the optimal cutoff points on the receiver operating characteristic curves. Model calibration was evaluated using Hosmer-Lemeshow goodness-of-fit (GOF) test (the R hoslem.test function) in the validation cohort, and the R plotCalibration function was used to plot the GOF calibration. A model was also fit after categorizing age (0-29, 30-59, 60-79, > = 80; years) and median household income (< 60, 60-80, > = 80; $1000). The beta coefficients of this model were used to design a severity heatmap. In order to enhance readability of the heatmap, risk scores were scaled to the minimum change in the coefficients. The p-values of the test of trend were reported in the derivation cohort. Because of the differences in testing criteria before and after April 29, 2020, a sensitivity analysis was conducted after dividing patients based on their corresponding reference dates. The same procedure as the main model were applied to the derivation and validation cohorts among patients tested before and after April 29, 2020. Additional sensitivity analysis was conducted on the population without discarding the 24 patients who have been hospitalized after the 30 days interval. In this analysis, these patients were considered as outpatients and a GLM was derived and examined.

The optimal cutoff for predicted probabilities was used to categorize patients into high risk category. Patients with estimated risks less than the above cutoff were then analyzed to calculate another optimal cutoff to define an intermediate risk category. Patients with estimated risk less than the second cutoff were reported as low risk. The same procedure was followed to group mortality risks of the hospitalized patients into low, intermediate, and high-risk groups. A Python implementation of the risk prediction model with categorized age and income is hosted at our website for seamless public access (https://dashti.bwh.harvard.edu/sars2/).

## Data Availability

The datasets analyzed during the current study are not publicly available. Due to patient privacy concerns, the underlying EHR data are not redistributable to researchers other than those prespecified in the approved Institutional Review Board application.

## Acknowledgments

We are grateful for the constructive comments from Dr. Nancy R. Cook, Brigham and Woman’s Hospital and Harvard Medical School. Authors are grateful for the support from the Enterprise Data Warehouse, Research Patient Data Repository, and COVID-19 Data Mart personnel at Mass General Brigham, in particular continuous helps from Stacey A. Duey and Julie M. Fiskio. This work was supported in part by the National Heart Lung and Blood Institute (T32 HL007575 to H.D., K24 HL136852 and HL 117861 to S.M., and 5K01HL135342 to O.D.), by 17IGMV33860009 from the American Heart Association to O.D., by the BWH Lerner Junior Faculty Research Award to O.D., and by philanthropic support from the Brigham and Women’s Hospital COVID fund.

## Authors Contributions

H.D., S.M., and O.D. were involved in the planning, conceptualization, and design of the study. H.D., E.C.R., D.W.B., and O.D. conducted data acquisition procedures, and performed the analysis. H.D., S.M., and O.D. were involved in interpretation of the data and analysis, and preparations of the manuscript. All authors reviewed the manuscript.

